# Loss-of-function myeloperoxidase mutations are associated with increased neutrophil counts and pustular skin disease

**DOI:** 10.1101/2020.02.21.20026161

**Authors:** Marta Vergnano, Katarzyna Grys, Natashia Benzian-Olsson, Satveer K Mahil, Charlotte Chaloner, Ines A Barbosa, Suzannah August, A David Burden, Siew-Eng Choon, Hywel Cooper, Nick Reynolds, Shyamal Wahie, Richard B Warren, Andrew Wright, The APRICOT and PLUM study team, Ulrike Huffmeier, Patrick Baum, Sudha Visvanathan, Jonathan Barker, Catherine Smith, Francesca Capon

**Affiliations:** Department of Medical and Molecular Genetics, School of Basic and Biomedical Sciences, King’s College London, London, UK; St John’s Institute of Dermatology, School of Basic and Biomedical Sciences, London, UK; Poole Hospital NHS Foundation Trust, Poole, UK; Department of Dermatology, University of Glasgow, Glasgow, UK; Department of Dermatology, Sultanah Aminah Hospital, Clinical School Johor Bahru, Monash University, Malaysia; Portsmouth Dermatology Centre, St Marys Hospital, Portsmouth, UK; Institute of Cellular Medicine, Newcastle University, Newcastle upon Tyne, UK; Department of Dermatology, University Hospital of North Durham, Durham, UK; Dermatology Centre, Salford Royal NHS Foundation Trust, Manchester NIHR Biomedical Research Centre, University of Manchester, UK; Centre for Skin Sciences, St Lukes Hospital, Bradford, UK; Institute of Human Genetics, Friedrich-Alexander-Universität Erlangen-Nürnberg, Erlangen, Germany; Boehringer-Ingelheim International GmbH, Biberach, Germany; Boehringer-Ingelheim Pharmaceuticals, Ridgefield, Connecticut, USA

## Abstract

The identification of disease alleles underlying human autoinflammatory diseases can provide important insights into the mechanisms that maintain neutrophil homeostasis. Here, we focused on generalized pustular psoriasis (GPP), a potentially life-threatening disorder presenting with cutaneous and systemic neutrophilia. Following the whole exome sequencing of 19 unrelated cases, we identified one affected individual harbouring a homozygous splice-site mutation (c.2031-2A>C) in *MPO*. The same homozygous change was subsequently identified in a further subject suffering from acral pustular psoriasis, a disease phenotypically related to GPP.

*MPO* encodes myeloperoxidase, an essential component of neutrophil azurophil granules. Of interest, the c.2031-2A>C allele was previously described as a genetic determinant of myeloperoxidase deficiency (MPOD), a condition which can causes recurrent infections. Here, a systematic literature review identified four individuals suffering from MPOD and pustular skin disease, further strengthening the link between *MPO* and pustular inflammation.

A subsequent analysis of the UK Biobank cohort demonstrated that the c.2031-2A>C allele was associated with increased neutrophil abundance in the general population (*P*=5.1×10^−6^). The same applied to three further MPOD mutations for which genotype data was available, with two alleles generating p-values <10^−10^. Finally, treatment of healthy neutrophils with an MPO inhibitor reduced cell apoptosis, highlighting a mechanism whereby *MPO* mutations affect granulocyte numbers.

These findings identify *MPO* mutations as genetic determinants of pustular skin disease and neutrophil abundance. Given the recent interest in the development of MPO antagonists for the treatment of neurodegenerative disease, our results also suggest that the pro-inflammatory effects of these agents should be closely monitored.

---

A tight regulation of neutrophil numbers is crucial to innate immune homeostasis. As mature granulocytes do not divide, their accumulation depends on the balance between progenitor proliferation, release of differentiated cells into the bloodstream and clearance of ageing cells^1^. Given the difficulty of manipulating primary neutrophils, the mechanisms that regulate these processes have mostly been investigated in animal models. In this context, the genetic characterization of human autoinflammatory diseases can provide crucial insights into the pathways that maintain neutrophil homeostasis.

Here we focused our attention on generalised pustular psoriasis (GPP, [MIM: **#**614204]), a potentially life-threatening condition presenting with flares of neutrophilic skin inflammation (pustular eruptions), fever, increased production of acute phase reactants and neutrophilia. While disease alleles have been described in *IL36RN, AP1S3* and *CARD14*, the majority of affected individuals do not carry deleterious changes at these loci^2^.

To identify novel genetic determinants for GPP we undertook whole-exome sequencing in 19 unrelated patients of varying ethnicity (Table 1, Figure 1a). Given the severity of the condition and the lack of parent-offspring transmissions, we hypothesised the presence of recessive loss-of-function alleles. We therefore filtered the patient variant profiles to retain rare homozygous changes predicted to cause premature protein truncation. This identified six candidate mutations, each affecting a single individual (Table S2).

**Table 1:**
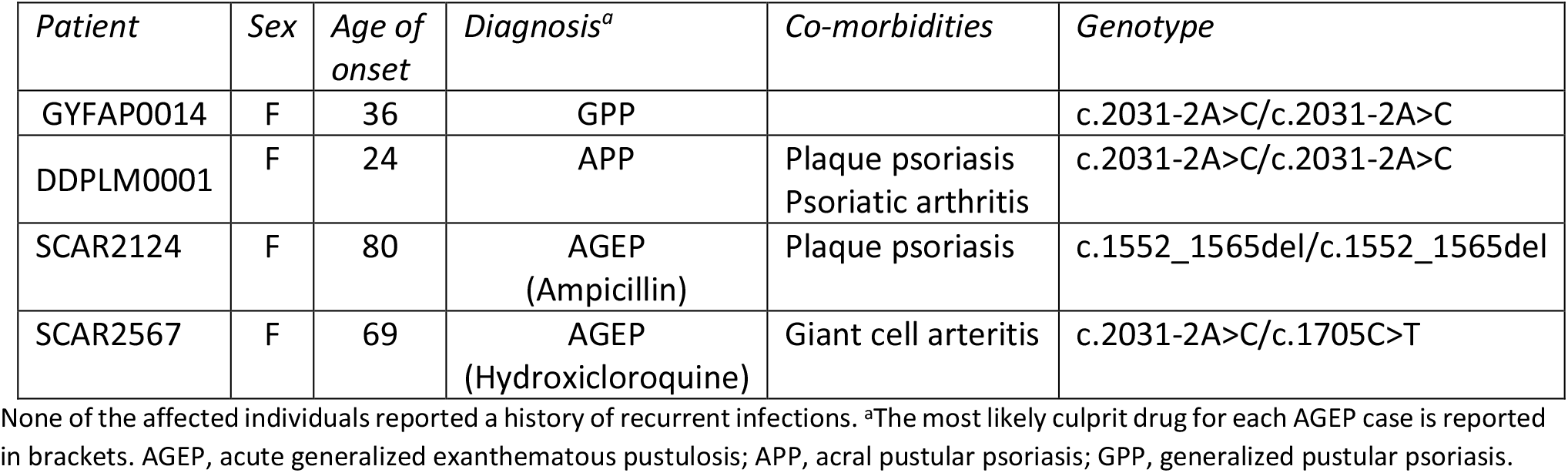
Features of the patients harbouring bi-allelic *MPO* mutations

**Figure 1:**
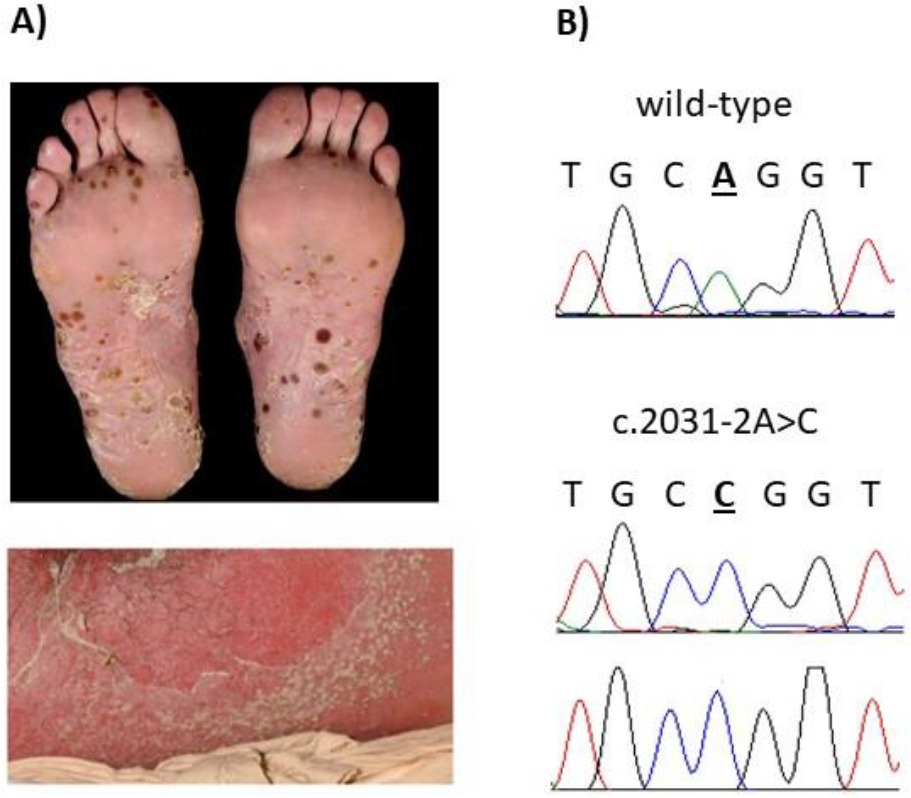
*MPO* mutations are associated with pustular skin disease. **A)** Typical presentation of generalised pustular psoriasis (bottom panel, showing skin pustulation on an erythematous background) and acral pustular psoriasis (top panel, showing neutrophil-filled pustules affecting the soles). **B)** Chromatograms showing the c.2031-2A>C substitution observed in the GPP and APP patient. The position of disease alleles is highlighted by bold, underlined font.

The c.2031-2A>C substitution in *MPO* (MIM: 606989) was selected for follow-up, as the gene encodes myeloperoxidase, a major component of neutrophil azurophilic granules. After the variant was validated by Sanger sequencing (Figure 1b), the entire *MPO* coding region was examined in 14 additional GPP cases and in 109 subjects suffering from acral variants of pustular psoriasis (APP). This uncovered a further affected individual harbouring a homozygous c.2031-2A>C substitution (Fig 1b). While no other bi-allelic *MPO* changes were detected, the frequency of the c.2031-2C/c.2031-2C genotype among European GPP/APP patients (2/142; 1.4%) was much higher than that observed in the controls sequenced by the gnomAD consortium (2/64,450;0.003%) (*P*=2.9×10^−5^). Of note, the c.2031-2C/c.2031-2C genotype was also absent from 590 British exomes processed with our in-house pipeline. Thus, the association with GPP/PPP is unlikely to be a technical artefact or to reflect population stratification between cases and controls.

Of note, the c.2031-2A>C change has been reported in individuals presenting with myeloperoxidase deficiency (MPOD, [#254600]), an inherited defect of neutrophil microbicidal activity^3^. Specifically, Marchetti et al demonstrated that the substitution affects splicing and leads to the production of a truncated protein lacking enzymatic activity^3^. Thus, this allele has a well-established impact on MPO function.

MPOD is a mild immune deficiency that is clinically well characterised. We therefore undertook a systematic literature review, to better understand the connection between *MPO* mutations, MPOD and skin pustulation. We examined 28 articles describing the presentation of MPOD in 217 individuals. This uncovered four cases where the disease manifested with pustular eruptions and a fifth where it was associated with the severe neutrophilic dermatosis known as pyoderma gangrenosum (Table 2). Given the very low prevalence of the above conditions (<1:100,000), these observations strengthen the link between MPO dysfunction and neutrophilic skin inflammation.

**Table 2:** Association of pustular skin disease and myeloperoxidase deficiency

| *Case report* | *Reference* |
| --- | --- |
| 46-year-old male with MPOD, plaque psoriasis and GPP.<br>Identical twin with mild pustular psoriasis. | Stendahl and Lindgren<sup>12</sup> |
| 61-year-old female with MPOD and annular pustular psoriasis | De Argila et al.<sup>13</sup> |
| 20-year-old male with MPOD developed generalized pustular eruptions following injury | Nguyen and Katner<sup>14</sup> |
| 81-year-old female with MPOD and pyoderma gangrenosum | Disdier et al.<sup>15</sup> |

GPP, Generalized pustular psoriasis; MPOD, myeloperoxidase deficiency

To investigate the mechanisms whereby *MPO* mutations contribute to disease, we explored the phenotypic effects of the c.2031-2A>C variant though a Phenome-Wide Association study (PheWAS). We queried the UK Biobank dataset, which includes genotype information and health data for a well-characterised population cohort (>450,000 individuals)^4^. A hypothesis-free analysis of 778 phenotypes revealed that the traits showing the most significant associations with c.2031-2A>C were related to leukocyte counts. In this context, the strongest effect size was observed for the association with neutrophil abundance (beta=0.45; *P*=5.1×10^−6^; Figure 2a). To validate these findings, we examined four additional MPOD alleles (p.Tyr173Cys, p.Met251Thr, p.Ala332Val, p.Arg569Trp) for which genotype data was available in UK Biobank. We found that all were associated with increased neutrophil accumulation, with p-values ranging from 0.008 to 3.9×10^−28^ (Figure 2b).

**Figure 2:**
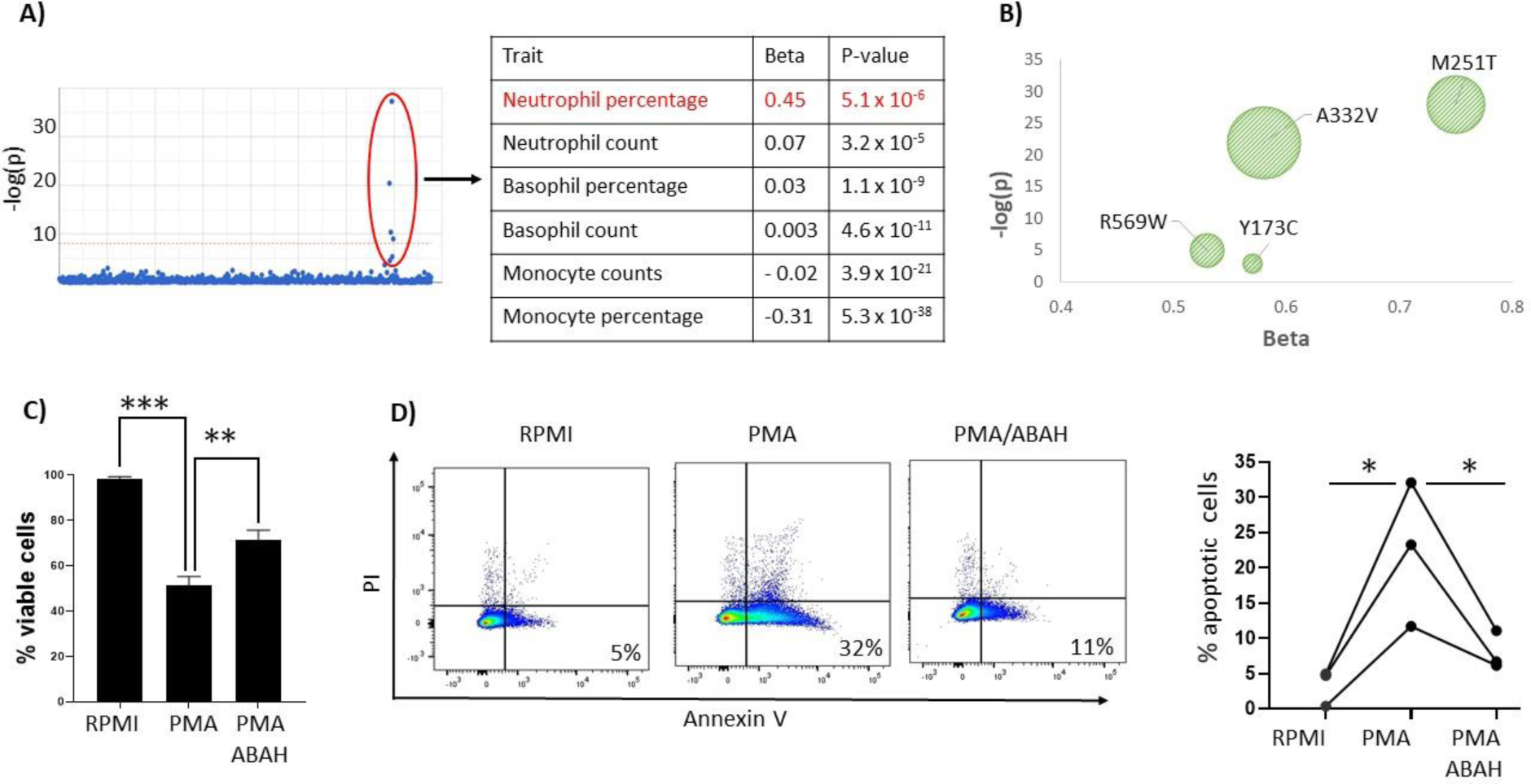
*MPO* mutations are associated with increased neutrophil counts and delayed apoptosis. **A)** Manhattan plot where each dot represents the association between c.2031-2A and a clinical trait. The p-values for phenotypes related to leukocyte counts are highlighted with a red circle and reported on the right, alongside the effect sizes (beta). **B)** Association between MPO deficiency alleles and neutrophil percentage. The size of each bubble represents the frequency of the mutation in UK Biobank. **C-D)** ABAH pre-treatment of cells stimulated with PMA increases viability (C) and down-regulates apoptosis (D). In C) Data are presented as the mean (+/-SD) of four experiments carried out in triplicate. In D) each line represents an independent healthy donor. A representative set of flow cytometry plots is shown on the left with the percentage of apoptotic cells (AnnexinV+, PE-population) for each condition. RPMI=untreated (medium only) *P<0.05 **P<0.01; ***P<0.001

To explore the pathways underlying the effects of *MPO* alleles on granulocyte numbers, we examined RNA-sequencing profiles generated in pure neutrophil populations (see Supplemental Methods). Specifically, we compared gene expression in the c.2031-2A>C homozygous GPP individual vs 11 healthy controls. We found that 95 genes were up-regulated in this subject (FDR<0.05) (Table S3). The majority of these loci (85/95) were not over-expressed in 7 unrelated cases (all *MPO* wild-type) examined in parallel, indicating that the changes are unlikely to be a secondary effect of inflammation. While the experiment was limited by the small sample size and the number of differentially expressed genes was too small for pathway enrichment analyses, we noted that two of the five most up-regulated loci (*PBK* and *GUCYA2*) encode proteins (PDZ binding kinase and soluble guanylate cyclase alpha-2 subunit) that can inhibit apoptosis^5; 6^. This suggests that *MPO* mutations may affect neutrophil survival.

We investigated this possibility by using a myeloperoxidase inhibitor (4-Aminobenzoic acid hydrazide, ABAH) to mimic the effects of *MPO* disease alleles in cell culture experiments. We induced neutrophil apoptosis through Phorbol 12-myristate 13-acetate (PMA) stimulation and assessed the effects of ABAH pre-treatment on this process. While PMA caused substantial neutrophil death, we found that ABAH supplementation caused an increase in cell viability (Figure 2c) and a reduction in the number of apoptotic cells (Figure 2d). Thus, neutrophil apoptosis is down-regulated in the absence of MPO activity.

Our findings demonstrate a significant association between *MPO* mutations and pustular skin disease. While the disease alleles described here have also been implicated in MPOD, we did not observe any evidence of immune deficiency in the individuals examined in this study. Likewise, pustular skin disease is only present in a fraction of MPOD cases. In this context, it is tempting to speculate that the manifestations of *MPO* mutations may be influenced by background polygenic variation. In fact, polygenic effects have recently been implicated in the modulation of rare, blood-related phenotypes^7^. The results of our PheWAS indicate that the effects of *MPO* alleles are likely to mediated by an up-regulation of neutrophil numbers. Of note, a significant association between granulocyte abundance and a common *MPO* variant has previously been documented^8^, further supporting the role of the gene in neutrophil homeostasis. While our PheWAS findings relate to circulating (rather than skin infiltrating) neutrophils, an increased prevalence of spondyloarthropathy has been reported among individuals with MPOD^9^, suggesting that MPO-related disruption of neutrophil apoptosis may affect multiple organs. Given that MPO inhibitors are being developed for the treatment of neurodegenerative disease^10^, our data suggest that the inflammatory side effects of these agents should be closely monitored during clinical trials.

Further experiments will be required to dissect the molecular mechanisms whereby MPO deficiency down-regulates cell death. Given that *PBK* (one of the most up-regulated genes in the c.2031-2A>C homozygous individual) is an inhibitor of myeloid cell apoptosis^5^, its role is worthy of further examination. A proposed link between MPO-related oxidative stress, NF-κB activation and apoptotic signalling^11^ should also be investigated. While experimentally demanding, these studies have the potential to illuminate key regulators of innate immune homeostasis and uncover new candidate genes for neutrophilic conditions.

## Data Availability

All relevant data is included in the manuscript

## Supplemental Data description

Supplemental Data include four tables.

## Consortia

### Membership of the PLUM and APRICOT study team

The following members of the PLUM and APRICOT study team contributed to this work: Thamir Abraham (Peterborough city Hospital),Mahmud Ali (Worthing Hospital),Suzannah August (Poole Hospital),David Baudry (Guy’s Hospital, London), Anthony Bewley (Whipps Cross University Hospital, London), Hywel Cooper (St Marys Hospital, Portsmouth), John Ingram (University Hospital of Wales), Susan Kelly (The Royal Shrewsbury Hospital), Mohsen Korshid (Basildon Hospital), Effie Ladoyanni (Russell’s Hall Hospital, Dudley), John McKenna (Leicester Royal Infirmary),Freya Meynell (Guy’s Hospital, London), Richard Parslew (Broadgreen Hospital, Liverpool), Prakash Patel (Guy’s Hospital, London), Angela Pushparajah (Guy’s Hospital, London), Nick Reynolds (Newcastle Hospitals), Catherine Smith (Guy’s Hospital, London), Shyamal Wahie (University Hospital of North Durham and Darlington Memorial Hospital), Richard Warren (Salford Royal Infirmary), Andrew Wright (St Lukes Hospital, Bradford).

Additional patients were recruited by A David Burden (Glasgow Western Infirmary), Siew-Eng Choon (Johor Bahru Hospital, Malaysia), Brian Kirby (St Vincent University Hospital, Dublin, Ireland), Alexander Navarini (University Hospital Zurich, Switzerland) and Marieke Sieger (Radboud Medical Centre, Nijmegen, The Netherlands).

## Acknowledgements

This research has been conducted using the UK Biobank Resource. We acknowledge support from the Department of Health via a Biomedical Research Centre award to Guy’s and St Thomas’ NHS Foundation Trust in partnership with King’s College London and King’s College Hospital NHS Foundation Trust (guysbrc-2012-1). The APRICOT trial is funded by the Efficacy and Mechanism Evaluation Programme (grant EME 13/50/17). UH is funded by the DFG (CRC1181, Project A05). MV is supported by a Medical Research Council (MRC) PhD studentship, NBO by a NIHR pre-doctoral fellowship (NIHR300473) and SKM by an MRC Clinical Academic Research Partnership award (MR/T02383X/1). RBW is supported by the Manchester NIHR Biomedical Research Centre.

## Declaration of interests

FC and JNB have received funding from Boehringer-Ingelheim. PB and SV are Boehringer-Ingelheim employees.

## Web resources

CADD, https://cadd.gs.washington.edu/

EGA, https://www.ebi.ac.uk/ega/home

GeneAtlas, http://geneatlas.roslin.ed.ac.uk/

GnomAD server, https://gnomad.broadinstitute.org/

OMIM, https://omim.org/

PubMed, https://www.ncbi.nlm.nih.gov/pubmed/

